# Impact of Mass Drug Administration with Ivermectin, Diethylcarbamazine, and Albendazole in Elimination of Lymphatic Filariasis in Five Districts of Nepal

**DOI:** 10.1101/2025.06.03.25328869

**Authors:** Ram Kumar Mahato, Gokarna Dahal, Yadu Chandra Ghimire, Rudra Prasad Marasini, David T.S Hayman, Kaliannagounder Krishnamoorthy, Sunil Raj Sharma, Dipak Sah, Ram Balak Ray, Radha Subedi, Keshav Raj Pandit, Sudip Raj Khatiwada, Achut Babu Ojha, Saroj Mahaseth, Deepak Bahadur Mahata, Molly Brady, Clara R. Burgert-Brucker, Briana Stone, Ashna Parajuli, Satya Raj Paudel, Bhim Prakash Devkota, Chandra Bhal Jha, Krishna Prasad Paudel, Bhim Prasad Sapkota, Bijay Bajracharya

## Abstract

**Background:** Nepal aims to eliminate Lymphatic Filariasis (LF) by 2030. Mass drug administration (MDA) has ceased in 53 of 64 endemic districts. In 2023, five districts with persistent LF (≥2% antigen prevalence) completed two rounds of MDA using a three-drug regimen (Ivermectin, Diethylcarbamazine, and Albendazole; IDA), achieving over 65% coverage. An Epidemiological Monitoring Survey (EMS) was conducted to evaluate IDA’s impact.

**Methods:** A cross-sectional EMS was conducted 9 months post-MDA in 11 evaluation units (EUs) across five districts, using two sites per EU (n=22). A total of 6,829 individuals aged ≥20 years were sampled via multi-stage methods, with ≥300 blood samples per site. Data on demographics and MDA participation were collected. LF antigen testing was followed by night blood microfilariae testing in antigen-positive samples. Analysis included non-parametric tests, logistic and mixed-effects models accounting for site-level clustering, and penalized regression (lasso and ridge) to assess predictor importance and manage multicollinearity.

**Results:** Nine of 11 EUs passed EMS. Two EUs in Kapilvastu failed due to ≥1% microfilariae prevalence in at least one site. Microfilariae prevalence was negatively correlated with site MDA coverage (p = 0.04), but not antigen prevalence (p = 0.8). Overall, 4.63% of participants were antigen-positive and 0.34% were microfilariae-positive (ratio 14:1). Being female (OR 0.12; 95% CI: 0.04–0.36) and participation in latest MDA round (OR 0.34; 95% CI: 0.15–0.77) were associated with lower microfilariae prevalence.

**Conclusion:** Nine EUs met the EMS threshold for impact assessment eligibility. Female gender and participation in the most recent MDA round were protective against microfilariae. Targeted MDA strategies focusing on men and high-risk areas are recommended.

## Background

Lymphatic Filariasis (LF), a mosquito borne filarial parasitic infection, causes disease conditions such as lymphedema and hydrocele. This disease is recognized globally as a neglected tropical disease (NTD) and remains a major public health problem in many tropical and subtropical countries and is the second leading infectious cause of disability worldwide (1). LF is targeted for elimination as a public health problem by 2030 using two key strategies: mass drug administration (MDA) with anti-filarial drugs to interrupt transmission and morbidity management and disability prevention (MMDP) to alleviate sufferings among those affected (2). In 2023, about 657 million people in 39 countries, including Nepal, have been estimated to live in areas that still require MDA to stop infection spread (3).

The national LF Elimination Program of Nepal has realigned the initial target of achieving LF elimination by 2020 (4,5) with the WHO’s NTDs Roadmap to meet the elimination target by 2030 (4,5). Out of 77 districts in Nepal, 64 are endemic for LF with approximately 27.7 million people (95.1% of the total population) at risk of infection at the beginning of the program (6,7). *Wuchereria bancrofti* transmitted by *Culex quinquefasciatus* is the only infection reported in Nepal (8). The Epidemiology and Disease Control Division (EDCD) under the Ministry of Health and Population (MOHP) has implemented the LF elimination program since 2003, initially using a two-drug regimen (diethylcarbamazine and albendazole, DA)(9). As a first step to determine how MDA has affected LF prevalence, WHO recommends sentinel and spot-check site surveys in an evaluation unit (EU); if prevalence is lower than 2% for LF antigen or 1% microfilariae in each site, the EU can progress to implementing a Transmission Assessment Survey (TAS; TAS-1). TASs are population-based cluster antigen surveys of 6-7-year-olds to determine if prevalence is low enough to stop MDA; a TAS should then be repeated two (e.g., TAS-2) and four (e.g., TAS-3) years after stopping MDA (10). By the end of 2023, MDA was stopped in 53 Nepali districts following successful demonstration of TAS-1. Of these, 47 districts have passed TAS-2 and 28 districts passed TAS-3. In 2022, a triple drug-regimen (ivermectin, diethylcarbamazine and albendazole; IDA) (11) was introduced in five districts (Morang, Kapilvastu, Dang, Banke and Kailali) that repeatedly failed to meet the ‘Pre-TAS’ standards (see below). This regimen was scaled up to 11 districts in 2023 as 10 districts failed Pre-TAS/TAS requirements, and one a newly classified endemic district (6,11).

As per the WHO’s provisional guidance on the triple drug regimen, an Epidemiological Monitoring Survey (EMS-previously, known as Pre-TAS) should be conducted 9 months after the last round of MDA following two effective (>65% epidemiological coverage at district level) rounds of MDA. The new survey requires the detection of microfilariae through night blood smears of all antigen positives, which differs from the previous practice of detecting only antigen positives. Districts and/or EUs that pass the EMS become eligible for the IDA Impact Survey (IIS-equivalent to TAS). Following the successful IIS, two additional IIS (similar to TAS-2 and TAS-3) should be conducted with a gap of two years post-MDA surveillance to ensure that infection levels remain below the thresholds (12). In 2023, an EMS was conducted in 11 EUs following two rounds of IDA in Morang, Kapilvastu, Dang, Banke, Kailali to assess whether the prevalence of LF had fallen below the transmission threshold of <1% microfilaremia and to identify key predictors of this outcome.

## Methods

The cross-sectional EMS was conducted using a multi-stage sampling design to determine whether two rounds of IDA MDA could reduce LF infection prevalence to below the transmission threshold, defined as <1% microfilariae prevalence, in sites within the EUs of 5 districts: Morang, Kapilbastu, Dang, Banke, and Kailali. Secondary objectives included evaluating MDA coverage, identifying predictors of LF infection, and comparing the ratio of LF antigen prevalence to microfilariae prevalence. The survey was carried out after 9 months of completion of MDA campaign and between 1^st^ January-30^th^ January 2024.

Stage 1: Five districts were selected where two rounds of IDA MDA had been completed with an effective epidemiological coverage of ≥65% at district level (MDA coverage for each district was determined from final MDA reports from EDCD).

Stage 2: The selected districts were divided into EUs, each with a population of not more than 500,000, as per WHO provisional guidelines (7,12). The EUs were defined based on risk factors, including low MDA coverage in the previous years, high number of resistant populations (i.e., populations that are reluctant to be medicated, high numbers of reported clinical cases, or proximity to low coverage areas. EDCD, in close coordination with partners and district program managers, established two EUs in Kapilbastu, Dang, Banke, and Kailali and 3 EUs in Morang (7).

Stage 3: EUs were the survey area and for sampling, two sites were selected – usually a sentinel site and a spot-check site. If a sentinel site did not exist or the sentinel site had <2% antigen prevalence in previous surveys, an extra spot-check site was chosen instead. If a spot-check site had an antigen level ≥2% in a previous survey it was included in this survey; otherwise, a new site was chosen. Ward, the lowest administrative unit in Nepal, served as the site. The sites were selected based on factors such as having high number of clinical cases, vector abundance, and poor epidemiological coverage of IDA MDA, with the assumption that if the most challenging sites had infection prevalence below the transmission threshold, the rest of the EU would also maintain a similar level. The sites selected in the 11 evaluation units and 5 districts are shown in Table S1.

Stage 4: A total of approximately 300 samples per site, or 600 samples per EU, were required, amounting to around 6,600 samples across 11 EUs. This sample size was calculated to detect a critical threshold of <2% with a 5% chance of Type I error and ∼75% power when the true prevalence is 1%, accounting for a design effect of 1.5 for populations under 2,800 or 2.0 for populations over 2,800. In total, 6,829 samples were collected to assess the impact of the LF IDA MDA.

The required number of households to achieve a sample size of 300 adults (population above ≥20 years) per site was calculated, accounting for an expected 25% non-response rate, and an average of 2.65 adults per household (7). This resulted in the selection of approximately 151 households per site, rounded to the nearest multiple of 50 for determining the segment size. During a ward-level meeting before the field survey, each site was divided into equal segments containing approximately 150 households and one segment was randomly selected from each site. All households within the selected segments were visited, and all available adults aged ≥ 20 years were asked to participate in the survey from a team of enumerators with the help of local health workers and female community health volunteers. Each eligible adult visited a temporary laboratory established nearby and were enrolled and interviewed about participation in the last MDA (did the person participate in the MDA and did they take the IDA) and whether they had ever been treated and tested for LF antigen using a rapid antigen test, Filariasis Test Strip (FTS). All antigen positives were then followed up on same day or following day for night blood collection, described below. Summary and detailed information on the EUs is in Tables S1-S4.

### Diagnostic Tests

The EMS employed FTS for antigen detection, followed by microfilariae testing for those who tested antigen-positive, to assess any potential transmission signals.

### FTS Testing

The FTS was used as the primary diagnostic tool to detect LF antigenaemia among the survey population. Blood samples were collected using a 75-μL blood collection pipette after pricking the left ring finger. The blood sample was then transferred to the sample application pad with the pipette, and the result was assessed exactly at 10 minutes. Each FTS was labeled with a unique ID so that tests could be linked with demographic information and other variables collected. FTS Test scores were recorded as negative if no test line was visible (but with a positive control line); and as positive if the test line was present (along with the positive control line). Tests with no control line or other issues such as no migration of blood were considered invalid results (13). In this situation, the test was repeated. All individuals who tested positive with FTS were further tested for microfilariae through night blood slide collection.

### Microfilariae Testing

Microfilariae testing was conducted using microscopy to detect microfilariae among all antigen positive individuals (14). The microfilariae testing was done either on the same night or the following night after the FTS results were confirmed. Trained lab staff were responsible for preparing one quality blood smear per person (slides), storing, and transporting them to laboratory facilities. The same staff member dried it for 24 hours, stained it properly using Giemsa solution, and read the slides under the microscope for the presence of microfilaria.

#### Sample collection

Night blood slides were collected between 10:00 PM and 2:00 AM to cover the peak appearance of microfilariae in the peripheral blood (15). After pricking the finger on the side of the finger pad, 60-μL of blood was collected into a calibrated capillary tube and applied in three parallel lines (approximately 20 µL each) along the length of the microscope slide.

#### Slide preparation and staining

The slides were air-dried overnight and then placed in distilled water for 5 minutes to dehemoglobinize the blood following standard procedures (10). After air drying for 1 hour, the slides were fixed in methanol for 5 minutes and stained with a 1:50 dilution of Giemsa stock for 50 minutes. The slides were air-dried again before being read under a microscope using 10X and 40X lenses to detect microfilaria.

#### Quality Control

All microfilariae-positive slides and negative slides were re-read by a second reader. The second reader was from the agency responsible for surveys within the MOHP having ‘Master Training of Trainers’ for LF microscopy and intensive experience of microfilariae microscopy to ensure accuracy.

### Data analysis

Prevalence of those antigen-positive and those microfilariae-positive were calculated for each site. For antigen, number of antigen-positive participants was divided by the number of antigen-positive and antigen-negative participants tested in the site. For microfilariae, the number of microfilariae-positive participants was divided by the sum of the microfilariae-positive participants plus microfilariae-negative participants plus antigen-negative participants (who were assumed to be microfilariae negative).

Differences in age distributions between antigen and microfilariae positive and test negative individuals were assessed using t-tests with Welch’s correction or Wilcoxon rank-sum tests where appropriate using R (v4.3.2). A one-sample non-parametric tests using IBM SPPS statistics 25 was employed to calculate 95% CI for proportions of samples positive for antigen and microfilariae. The samples were grouped by age class (20-29, 30-39, 40-49 and 50 and above) and analyzed antigen and microfilariae positives. Percentages were calculated to compare both 1) the positivity rates of LF antigenemia and microfilaremia out of the total number of people sampled, and 2) the percentage of microfilariae test positive out of those antigen positives. Correlation analyses, Chi-square tests (χ^2^) and odds ratios (OR) were used to assess the associated factors with MDA coverage.

Binary logistic regression was applied to identify the significance of three hypothesize predictors of microfilariae positivity – age, gender and treatment in last MDA. Because of the study design with multiple sites and two replicates within those sites, the data are clustered—i.e., observations within a site are likely to be more similar to each other than observations from different sites. This violation of the independent observations assumption meant we also used a multi-level model to account for this clustering by allowing for random effects at the site level, which captures the variation between sites. Here, we used a Generalized linear mixed model (glmm) fit by maximum likelihood (Laplace Approximation) from the R *lme4* package with the same age, gender and MDA treatment predictors.

We used logistic regression to account for confounders (e.g., age, district) and assess interactions between four hypothesized predictors – gender, age, district, and treatment in the last round of MDA. In these analyses, age was used as a continuous variable in contrast to above. For the multivariate regression, we first attempted standard binomial logistic generalized linear regression, starting with all interactions among the four potential predictors and using glmm with district as a random effect. Preliminary analyses using standard binomial logistic generalized linear regression and model choice by AIC in a stepwise algorithm suggested all these variables might be important. However, since there were few microfilariae positive cases and low positive case numbers failed to find interactions significant, despite being in the best model by AIC, we fit a logistic generalized linear model using the same four predictor variables via penalized maximum likelihood using both Lasso and ridge regression in the *glmnet* R package (16). The data used in these models is plotted by the predictor variables in Figures S1 and S2.

## Results

The survey involved 6829 participants aged 20 years and above. All data are shown in Figures S1 and S2 by antigen and microfilariae status respectively.

Site-level coverage of ≥ 65% was evident in the most recent round of IDA MDA across several sites in the EUs (Table S3). These sites successfully passed the EMS as the microfilariae prevalence was found to be <1%. Coverage of ≥80% was observed in Dangisharan and Sisania (Dang B), Salyanbagh and Narainapur (Banke A), Baijapur and Rajhena (Banke B), Pahalmanpur (Kaialali A) and Janaki (Kaiali B). Except for Dangisharan (Dang B), all these sites recorded 0% microfilariae positivity. Sites such as Sisaniya (Dang B), Salyanbagh and Narainapur (Banke), Baijapur Rajhena (Banke) and Pahalmanpur (Kailali A), which had ≥90% coverage in the recent round of MDA, also recorded 0% microfilariae positivity. Some sites such as Sundarharaicha (Morang A), Patharisanischare (Morang B), Bakharitol (Morang C), and Dhangadhi (Kailali), passed the EMS despite having <65% coverage in the recent round (Table 1). Some sites like Shivraj (Kapilvastu A) and Bahadurgunj (Kapilvastu B) had microfilariae prevalence ≥1% with MDA coverage <65% and thus these sites failed EMS.

**Table 1:**
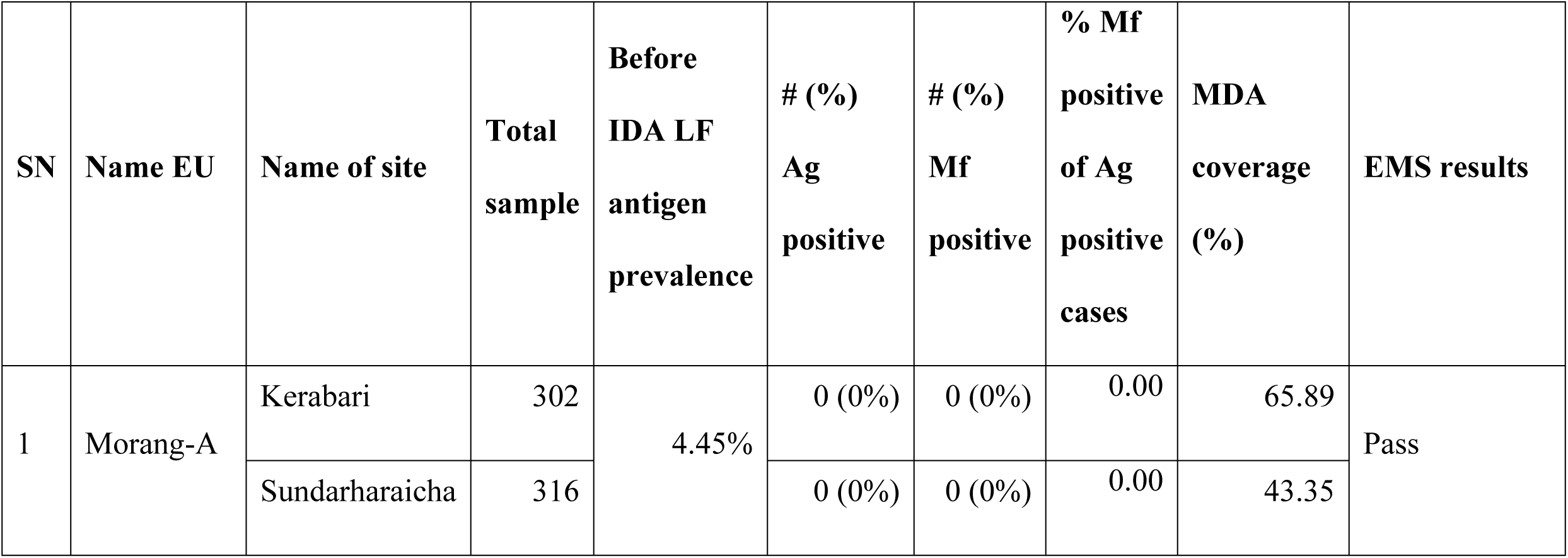
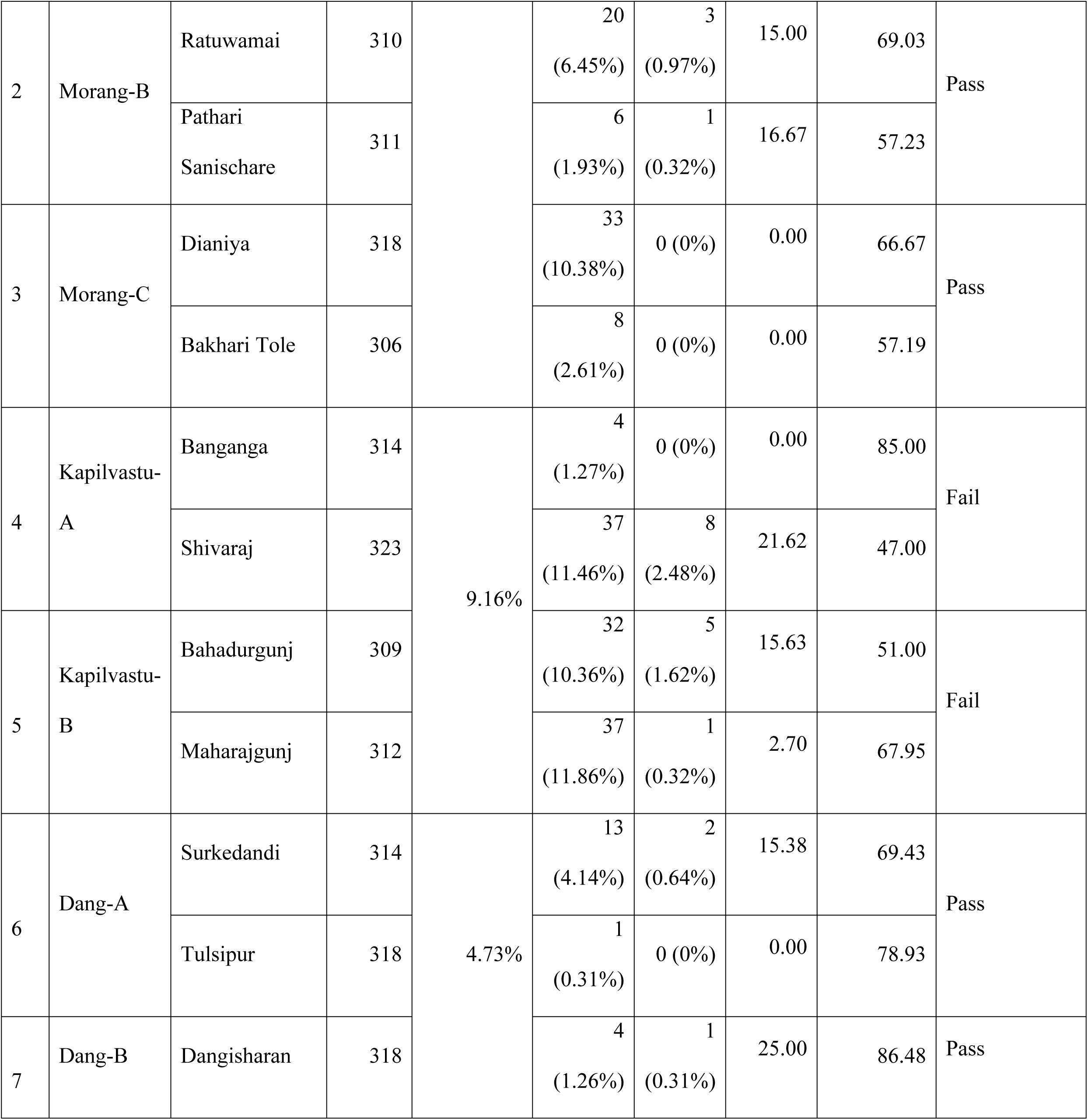
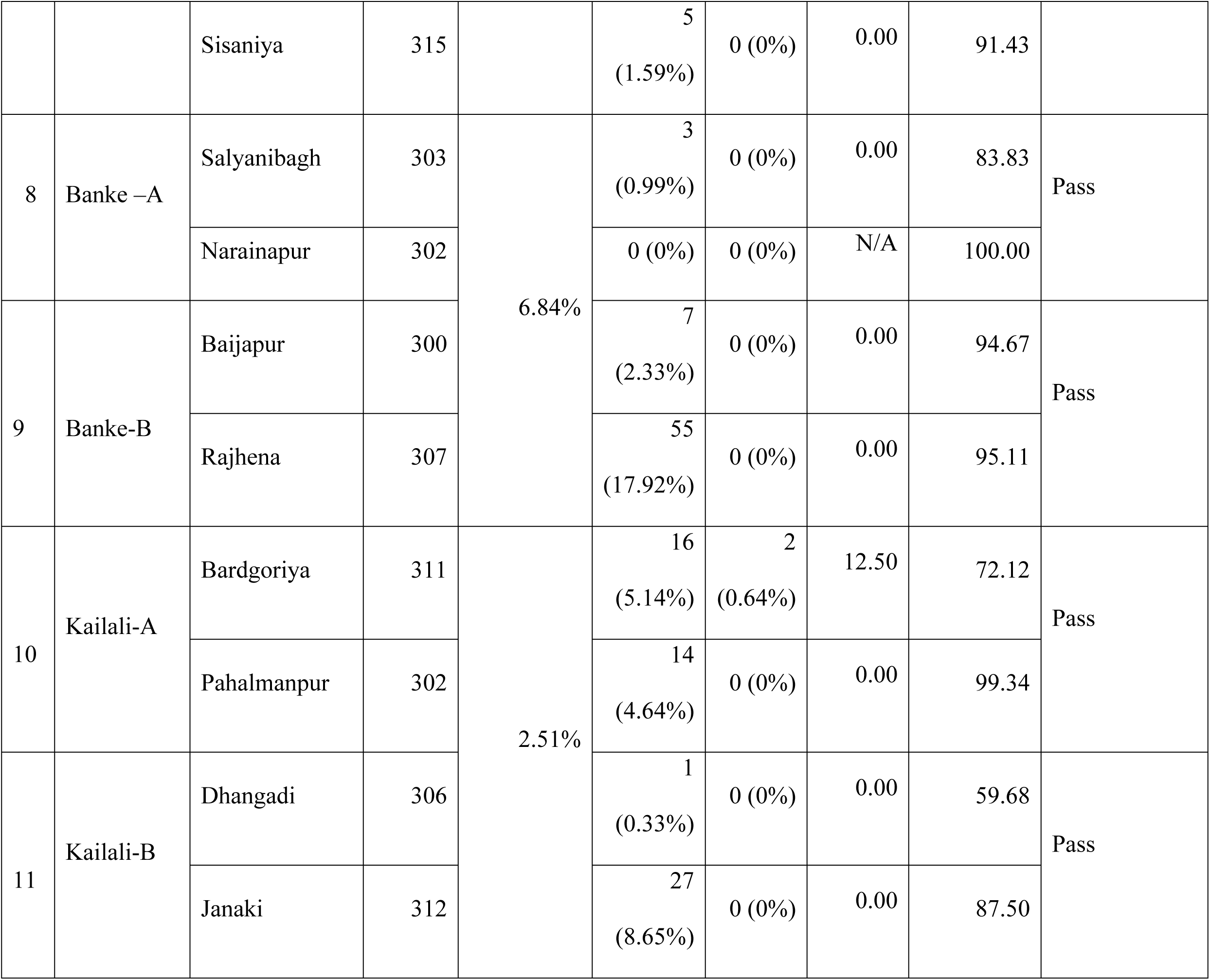
Site-specific antigen (Ag) and microfilariae (Mf) prevalence and mass drug administration (MDA) coverage in the last round.

Overall, 9 out of 11 EUs across the 5 districts passed EMS after two rounds of IDA MDA, indicating infection prevalence below the transmission threshold. Despite LF antigenemia prevalence being ≥2% in 9 sites, microfilariae prevalence remained <1% with IDA MDA coverage ≥65%. Bakharitol, although showing <1% microfilariae and ≥ 2% LF antigenemia, had inadequate coverage of LF MDA. A negative correlation was found between epidemiological coverage in the last round of MDA and microfilariae prevalence (τ = -0.35, p-value 0.04) in the 22 sites, but no relationship was found between the coverage in the last round of MDA and antigen prevalence (τ = -0.035, p-value 0.8) (see Figures S3 and S4). Overall microfilariae positivity was 7.12% among LF antigen-positive individuals and the ratio of antigen-positivity to microfilariae-positivity was 14:1.

There was an overrepresentation of women participants in the sample, with 67.1% being female. The proportion of males was lower across all age groups compared to females (Figure S5; Table S5). The median age was 40 years (IQR: 30–55). The overall mean age of the participants was 43, whereas the mean age of individuals who tested positive for LF antigen was significantly higher by approximately 3 years compared to those testing LF antigen negative, with a mean age of 46 (t = -3.2, df = 343, p value = 0.002), and those microfilaria positive were higher still at 49 years (Table S6), but not significantly different to those testing negative for microfilariae (Figure S6; t = -0.72, df = 24, p-value = 0.48). These results were consistent with those using Wilcoxon rank sum test with continuity correction. Prevalence by age class (e.g., 20-29, etc.) were not significantly different, but age class prevalence estimates had wide confidence intervals (Figures S6 and S7; Table S5). Age was not a significant predictor in binary logistic regression (Table S8; p-value 0.4).

The survey found females were 50% less likely to report having never been treated compared to males (OR 0.50; 95% CI 0.42-0.59) (Table S7). At the same time, females were 33% less likely to report non-treatment in the most recent round compared to males (OR 0.67; 95% CI 0.6-0.75) (Table S8).

Using univariate analyses, we found that gender (OR 0.12; 95% CI 0.04-0.36) and treatment in the most recent MDA round (OR 0.37; 95% CI 0.15-0.77) were significantly associated with reduced microfilariae rates in the community (Table S9, Figure 1).

**Figure 1.**
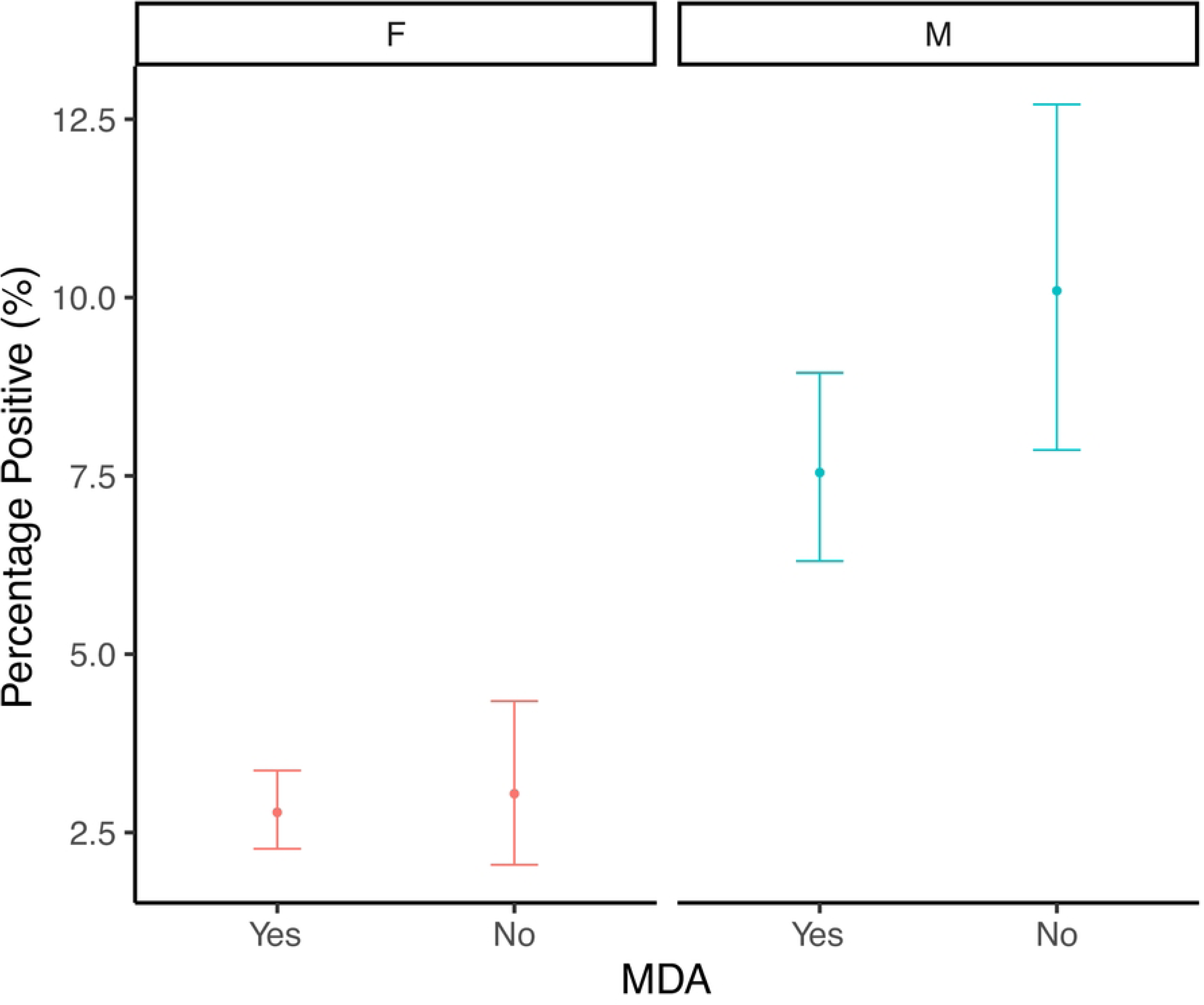

Being male was the only significant predictor (β = 1.13, p-value <0.05) in the multi-level *glmm* with random district effects, but with a p-value close to 0.05 (0.048). Neither age (β = 0.007, p-value 0.6) nor MDA treatment (β = -0.81, p-value 0.08) was significant.

Multivariate analyses, however, suggested there were multiple interactions as the best models by AIC included all covariates, including districts. Ridge and Lasso regressions showed that being male and living in Kapilvastu were the most important predictors of being *antigen* positive. Being treated in MDA was the most important protective factor, along with relatively lower rates in Morang and Dang districts (Figure S8). However, for *microfilariae* infection (Figure 2), the most important relative predictors were being from Kapilvastu and Dang and being male (Figure S9). Those treated in MDA were 66% less likely to be microfilaria positive (OR 0.34, 95% CI 0.15-0.77), and being treated in MDA was the most important protective factor. Age was not important in either analysis accounting for these factors. However, the relative importance of participants being from Kailali switched between analyses (Figures 2).

**Figure 2.**
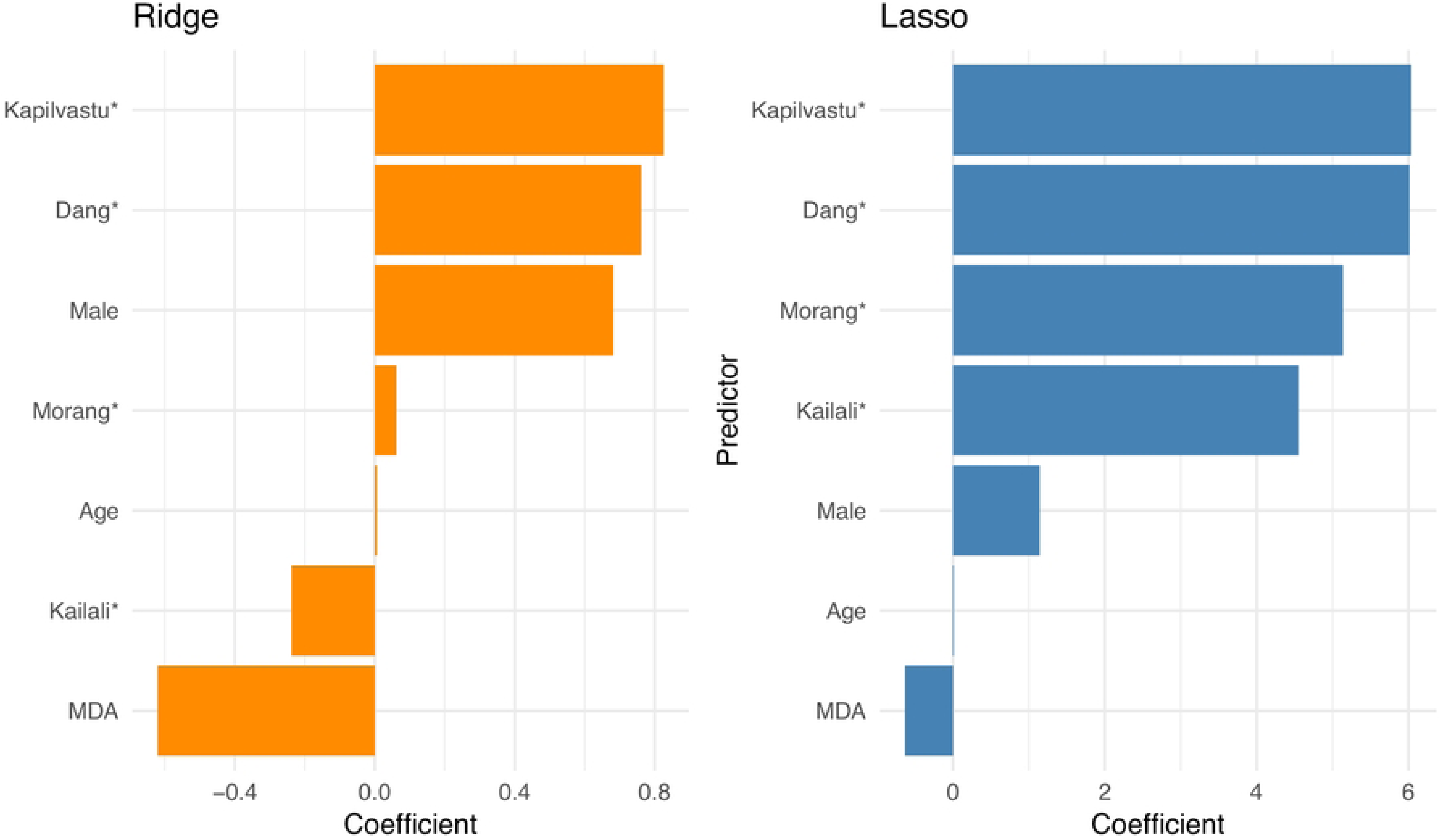

## Discussion

We conducted a cross-sectional, community-based EMS to assess the prevalence of LF antigenemia and microfilaremia to assess the impact of the IDA regimen in the districts. The survey involved 6,829 participants aged 20 years and older. The WHO’s New M&E Guidance on Monitoring and Epidemiological Assessment of Mass Drug Administration identifies that the age group ≥ 20 years carries the highest microfilariae burden and represents the greatest risk for propagating LF in the community. The selection of this target age group is also consistent with previous work (17–19). The guidance also identifies that a lack of microfilariae in adults is a good indicator that there is no ongoing transmission of LF in the community.

Five (23%) out of 22 sites from 11 EUs showed less than 65% LF MDA coverage, a target coverage at implementation unit level for LF elimination. The remaining sites all met the target coverage (10,12), with nine (41%), and five sites (23%) achieving ≥ 80% and ≥ 90% MDA coverage respectively. The two EUs which failed EMS had higher LF antigen prevalence in 2022, i.e., before starting 2 rounds of IDA, and less than 65% LF MDA coverage at site-level in the recent round of MDA. The prevalence of microfilaremia is statistically associated with LF MDA treatment in recent round, with an odds ratio of 0.34 (95% CI 0.15-0.77), showing the impact of MDA comparable with previous studies (19–22). These results highlight the need for ongoing work to strengthen MDA to eliminate transmission, particularly in sites with higher transmission.

A field trial in India with IDA reported 84% clearance of microfilariae (23). The protective factor with the recent round of IDA MDA in our study was found to be, less than some previous studies (20,24,25), but comparable with others such as Côte d’Ivoire in 2019 (26). Therefore, given the association with IDA and microfilaria positivity, and the Lasso and ridge regression results, the infection persistence in the EUs with evidence of continued transmission despite IDA MDA is likely due to suboptimal coverage (<65%).

Overall microfilariae positivity was 7.12% among LF antigen-positive adults and the ratio of antigen-positive to microfilariae-positivity was found to be 14:1. Similar results are reported in previous studies (19,28). A single worm or worms of either sex that do not produce microfilaria (28–30) can infect people and taking MDA may clear the microfilaria within one month, yet the antigen may persist for years (31), so the microfilariae-antigen ratios may reflect the parasite life cycle and the effects of treatment on microfilariae (32). This likely explains why the microfilariae prevalence was impacted by the last round of MDA (p-value 0.04), whereas the antigen prevalence was not (*p*-value 0.8). WHO assumes a ratio of 2:1 antigen-positivity to microfilariae-positivity for threshold values; further studies and meta-analysis of data could help determine a more appropriate ratio in post-MDA populations.

More generally, we found no correlation between age and microfilariae prevalence in contrast to other studies, such as in Egypt and Tanzania (18,19). If transmission has been ongoing and endemic for a long time, all adults may have had similar cumulative exposure to infected mosquitoes, leading to similar prevalence across age groups, though lower prevalence in older age classes may indicate reduced burden due to effective MDA.

We found a lower microfilariae prevalence in females compared to males (Figure 1; OR 0.12, 95% CI 0.04-0.36), and a similar result was found in Tanzania (19). Exposure to mosquito bites and different participation rates in MDA may have influenced the gender difference seen in this survey. We found that females had significantly fewer ‘never treated’ issues compared to males and in the recent round treatment issues were significantly lower in females compared to males, matching findings in other countries (19,33). During the survey, the ratio of males in all age groups was lower compared to females. These findings are comparable to other studies with the 7.5% absentee rate, among which 88% were males, and among the male absentees, more than 76% were between 15 and 34 years of age (34,35). Together, this suggests greater efforts are needed to find, test and/or treat men, as they are both more likely to be infected and less likely to participate in MDA and surveys.

The present survey clearly showed spatial heterogeneity in MDA treatment which require novel strategies to reduce the treatment gap in subsequent two additional MDA rounds in repeat MDAs in the EMS failed districts. It is also necessary to identify and address other potential barriers to participation, devise the most effective messages and channels for conveying health information, and devise effective drug administration strategies before undertaking the additional MDA rounds (36). High-risk groups (never treated and elderly) should be identified and targeted, as advocated by Lau et al., 2020, and others (37–39).

Lastly, despite the large sample size, the infection prevalence was low, making it challenging to analyze interactions among predictor covariates and potential confounders, including accounting for spatial clustering. Future analyses could better address multiple comparison issues (i.e., Type I error) and might benefit from the use of other multi-level models (such as hierarchical or mixed-effects models) to analyze additional data. These models would help avoid pseudo-replication and allow for more robust inferences about variation at different levels and interactions among variables. In this study, we used logistic regression without interactions (Table S9), as well as ridge and Lasso regression (Figure 2), to handle the large but sparse data matrices. However, other approaches, including Bayesian methods, could also be considered.

## Conclusion

The EMS conducted across 11 evaluation units in 5 Nepali districts revealed that 9 out of these 11 units achieved a microfilariae prevalence of less than 1%, suggesting that the infection prevalence is below the transmission threshold. The low prevalence of microfilariae is attributed to factors such as higher MDA coverage in the campaign and a higher proportion of female participants. LF MDA treatment was notably higher among females compared to males with correspondingly higher microfilariae prevalence among men, suggesting greater efforts may be needed to ensure men are treated in the future. Similarly, some locations have higher transmission and should be targeted. The recent triple-drug regimen demonstrated a significant reduction in the prevalence of LF, however, suggesting that the current programs are successful and have the ability to eliminate LF transmission.

## Data Availability

All Data are presented in the paper and if raw data are required, it is available upon request!

## Contributors

RKM conceived and designed the manuscript; RKM, GD, KRP, RPM, RBR, DS, RS, ABO, SRK, SM, DBM managed and supervised the data collection; RKM, DTSH analyzed the data; RKM, GD, BB, SRP wrote the original draft. YCG, CBJ, SRK, DTSH, BB, KK, SRS, AP, BPD, BPS, KPP, MB, CB, and BS reviewed and revised the manuscript. All authors read, reviewed, and approved the final manuscript.

## Data availability

The datasets generated and/or analyzed during the current survey are not publicly available to safeguard the privacy of individual patients. The data are available from the corresponding author upon reasonable request.

## Ethics approval, consent to participate, and consent for publication

This survey was carried out by EDCD and VBDRTC; both institutions are the major implementing bodies for management of vector borne diseases under Nepal’s Ministry of Health and Population (MoHP). The activities conducted by MoHP as a part of the set strategic goals for the regular monitoring and programmatic progress in National Lymphatic Filariasis Elimination Program (NLFE) have been exempted by Nepal Health Research Council (NHRC) from the ethical review process (**Ref. no 1530 NHRC, December 10, 2020**). Nevertheless, written informed consent was obtained from all the participants prior to interviews and blood sample collection.

